# Understanding the primary healthcare context in rural South and Southeast Asia: a village profiling study

**DOI:** 10.1101/2024.09.03.24313043

**Authors:** Rusheng Chew, Sazid Ibna Zaman, Mst. Asfat Ara Joly, Didar Uddin, Md Nurullah, James J Callery, Carlo Perrone, Thomas J Peto, Koukeo Phommasone, Aung Pyae Phyo, Wanlapa Roobsoong, Aninda Sen, Moul Vanna, Arjun Chandna, Tiengkham Pongvongsa, Lek Dysoley, Nicholas PJ Day, Yoel Lubell, Richard J Maude

## Abstract

The use of comprehensive village profiles is one way of characterising contextual factors important for the implementation of primary healthcare interventions and service planning in rural areas. However, there are few such data available at the village level in rural South and Southeast Asia. This study aimed to address this gap, as well as compare high-level data from representative under-served and understudied villages across seven sites in five countries (Thailand (n=3), Cambodia, Laos, Myanmar, and Bangladesh). A survey-based approach using key informants supplemented by other relevant information sources was used to collect data from 687 of 707 villages participating in the South and Southeast Asian Community-based Trials Network. Data on four key health and socio-economic indicators (literacy rate, percentage of attended deliveries, percentage of fully-immunised children, and percentage of latrine coverage) as well as access to health services, public utilities, and education were collected and analysed using descriptive statistics. There was considerable variation between sites in terms of health and socio-economic indicators given that the countries are at different stages of development, and also between the three sites in Thailand. Five of the seven sites were highly diverse ethno-culturally and linguistically, and all were reliant on primary health centres as well as village health workers/village malaria workers as the main providers of primary healthcare. These were generally bypassed by severely ill patients in favour of first-level referral hospitals and private sector facilities in towns. While >75% of villages at each site were near to a primary school, educational attainment was generally low. Over 70% of villages at each site had mobile phone coverage and availability of electricity was high (≥65% at all sites bar Myanmar). These results illustrate the wide diversity of villages in rural South and Southeast Asia that need to be considered in public health research and policymaking.

## Introduction

An understanding of contextual factors is crucial to the successful implementation of interventions to improve healthcare delivery. Although there is no one standard definition of ‘context’ as it relates to healthcare,[1] one useful definition is the “situational opportunities and constraints that affect the occurrence and meaning of organizational behaviour as well as functional relationships between variables”,[2] indicating that context necessarily includes both internal and external inter-related factors.[3]

In rural South and Southeast Asia, healthcare is principally delivered at the village, or village group, level through village health workers and primary health centres, with referral facilities typically being distant and relatively difficult to access.[4, 5] As such, interventions targeting primary healthcare would be expected to produce maximum impact. Nevertheless, as previous experience with primary health centres in India has shown, despite the apparent basing of health service provision on the principles of equity and administrative accountability, not catering to local contextual variations between villages and not involving local staff in service planning resulted in sub-optimal healthcare delivery.[6] The key variables identified relate to service coverage and socio-economic and geographical factors,[6] the determination of which have the benefit of being useful for micro-planning i.e., planning of health services and interventions at the village or sub-district level.[7]

Constructing concise yet comprehensive village profiles which contain relevant key indicators is one way of characterising contextual factors important for the implementation of primary healthcare interventions and service planning. Such indicators must be as simple as possible, widely accepted, and highly applicable to the village setting. They should also be easily comparable, sensitive to change, and yield data which is meaningful and easy to analyse.[8]

While governmental authorities may have village-level data on some of these indicators, these may not be available from a single source and may not be current, especially as access to rural and remote locations is difficult. Correspondingly, there is also scant published research which aimed to depict the rural South and Southeast Asian primary healthcare context via the profiling of representative villages, let alone research which aimed to compare high-level village profiling data across countries. This study aimed to fill these knowledge gaps, primarily to understand better the findings of a large-scale observational study on the epidemiology of acute febrile illness in the region in which patients were recruited from the selected villages,[9] but also to use the profiles to plan future research and guide policy-making.

## Methods

### Setting

This study was carried out across 687 of the 710 villages forming part of the South and Southeast Asian Community-based Trials Network (SEACTN), which spans seven sites across five countries (Thailand, Laos, Cambodia, Bangladesh, and Myanmar) (Figure 1). The sites are located in rural districts of the following locations: Chiang Rai, Tak, and Yala provinces, Thailand; Savannakhet province, Laos; Chattogram division, Bangladesh; Battambang and Pailin provinces, Cambodia; and Karen state, Myanmar. A full description of SEACTN is available in the open letter announcing its launch.[9] Every village included in SEACTN was profiled, with the exception of 20 villages in Tak province, Thailand, and three villages in Karen state, Myanmar which were not studied due to logistical difficulties and security concerns, respectively.

**Figure 1.**
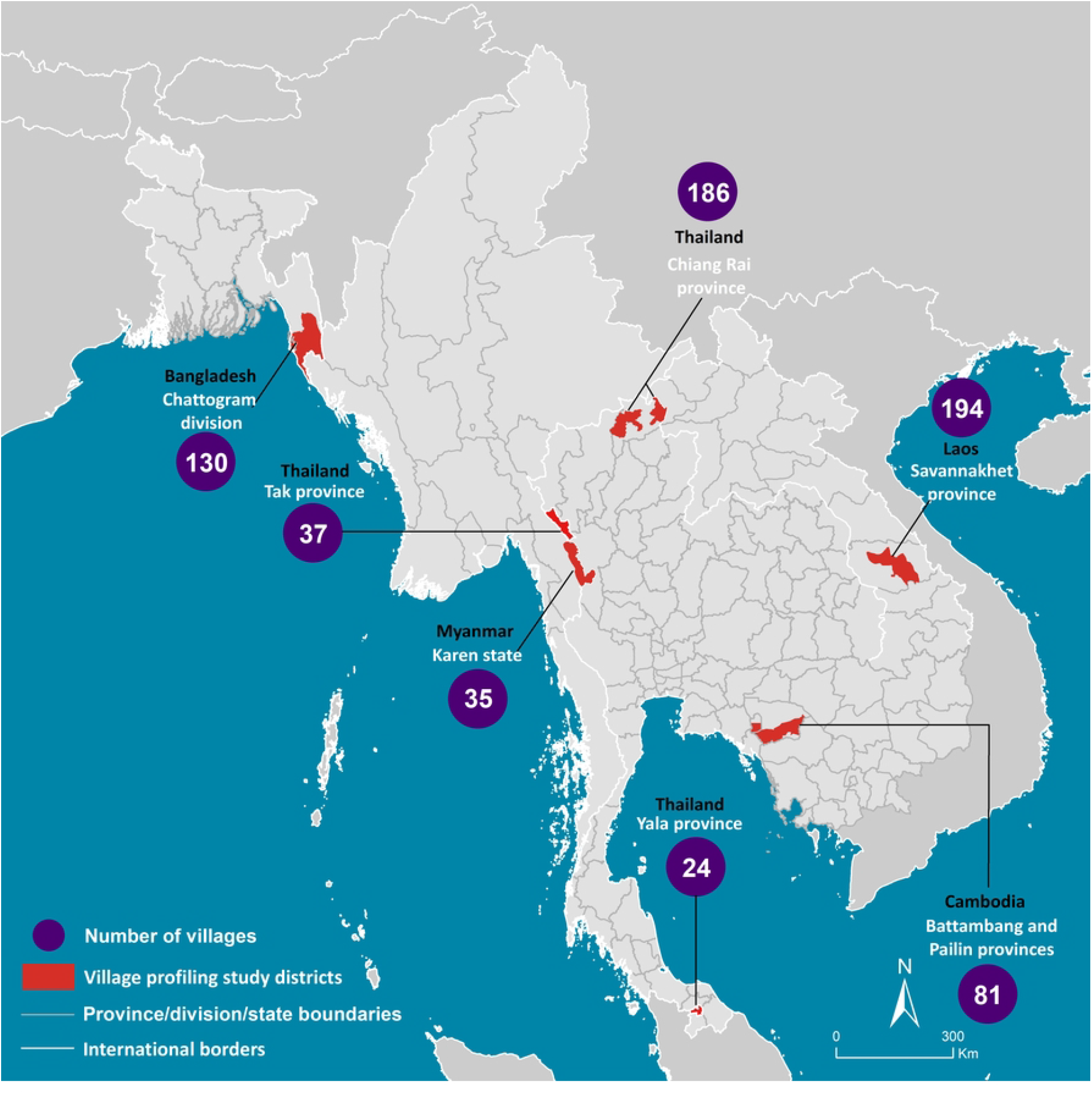
Locations of study sites in the South and Southeast Asian Community-based Trials Network and the number of villages profiled at each site.

Participating villages were selected for inclusion in SEACTN by country implementing partners, who have the most knowledge to determine which villages are representative of the areas where the target populations for the observational study and other SEACTN project components reside, and where prospective, directly-collected data would be most useful. Of note, one of the primary objectives of establishing SEACTN, which was to map and describe health services in under-served and under-studied rural and remote areas of South and Southeast Asia, was a guiding principle in selecting villages.

An exception was made for the Myanmar site due to the ongoing armed conflict in Karen state,[10] resulting in the need to reduce inaccessibility to ensure the viability of conducting other SEACTN activities. Therefore, while all villages profiled were rural, these were in the area near the Thai border rather than in the interior of Karen state, and better connected to transport and communication infrastructure. Because of this, and their borderland location, residents of these villages were also able to access facilities in Thailand, in particular health services.

### Study design

Similar to previous studies which collected data on key health indicators,[11] we adopted a survey-based study design, with key informants used as respondents. These key informants were identified by site research staff, and included healthcare workers, village leaders, and villagers who were judged to have the requisite knowledge. Other data sources were used as relevant; these included Global Positioning System (GPS) data, census data, and published government documents e.g., official statistics.

### Questionnaire development

The survey questionnaire was developed iteratively through a series of discussions with senior researchers from each site. This approach was taken to ensure that only relevant and useful data were collected and, for questions where respondents were presented with options from which to select, the options were tailored to site-specific conditions.

The questionnaire consisted of four sections covering village location, socio-economic conditions, health services and infrastructure, and public utilities. Questions on key health and socio-economic indicators were adapted from the indicators of resources and service performance identified by Larson and Mercer as being well-defined, valid, feasibly collected, and useful i.e., literacy rate, percentage of attended deliveries, percentage of fully-immunised children, and percentage of latrine coverage.[8]

Given country-specific differences in the definition of literacy, a proxy measure of completion of at least five years’ schooling in adults aged ≥18 years was used instead. As per the World Health Organization, an attended birth was defined as one presided over by a skilled birth attendant i.e., an accredited health professional, such as a midwife, doctor or nurse, who has been educated and trained to proficiency in the skills needed to manage normal uncomplicated pregnancies, childbirth and the immediate postnatal period, and in the identification, management and referral of women and neonates for complications.

Traditional birth attendants, whether trained or not, are not classed as skilled birth attendants.[12] A fully immunised child was defined as one who has received the following vaccines: Bacille Calmette-Guérin; three doses of diphtheria, pertussis, and tetanus; three doses of polio; and measles by the age of one year.[8] Estimates for educational attainment, skilled birth attendance, and vaccine coverage were obtained by asking key informants to consider a representative sample of 20 persons relevant to the question, thus are rounded to the nearest 5% at the point of data collection.

The WHO/United Nations Children’s Fund Joint Monitoring Program for Water Supply, Sanitation, and Hygiene (WHO/UNICEF JMP) defines ‘improved drinking water sources’ as those that are likely to be protected from outside contamination, particularly faeces. Examples include household connections, public standpipes, boreholes, and protected dug wells.[13] The WHO/UNICEF JMP also distinguishes between ‘improved’ and ‘unimproved sanitation facilities’, with the former defined as those that hygienically separate human waste from human contact including flush or pour-flush to piped sewer systems, septic tank pit latrines, ventilated-improved pit latrines, or pit latrines with slab or composting toilets. Importantly, shared or public-use sanitation facilities are not considered to be improved.[13] To describe access to health services and other relevant infrastructure and public utilities, key informants were asked to list the services available in the village or within 30 minutes’ walk of their residence. This 30-minute travel time was chosen as it has been widely used to define appropriate accessibility.[14, 15]

The survey questionnaire template, which formed the basis for the study electronic data collection tool described in the next section, is shown in Supporting Information S1.

### Data collection instrument and procedures

An electronic version of the survey questionnaire was developed using KoboToolbox (Kobo Organization, Cambridge, USA) and loaded onto tablet devices. Data collection was then carried out electronically for each village by site research staff using a combination of visits to the village, telephone interviews, and desk-based research. Village population figures were obtained from the latest available official statistics; for villages where these were not available, estimates were obtained through key informant interviews.

Key informants were interviewed either in person or by telephone. At sites where more than one key informant was interviewed per village, the mean of the responses to questions requiring numerical answers was entered as the final value if the responses were broadly similar. Otherwise, research staff were allowed to use their discretion in selecting the preferred response based on how well-suited the knowledge and expertise of the key informant was to the question e.g., the response of a health worker key informant would be preferred over that of a village head for the question on percentage of attended deliveries.

### Statistical analysis

Data were analysed and presented using descriptive statistics to depict an ‘average’ village for each site. Count data at the site level were reported directly, while medians and upper and lower quartiles were calculated for continuous data captured at the village level. Percentages were calculated for binary and categorical data captured at the village level.

Responses to questions requiring the selection of one or more responses e.g., types of water source available at each village, were summarised graphically to show the proportion of villages per site with access to a particular service. Analyses were carried out using Excel (Microsoft, Redmond, USA).

### Ethical approval

Ethical approval was not required for this study.

## Results

The locations of the villages profiled in this study within each study site are shown in Figure 2.

**Figure 2.**
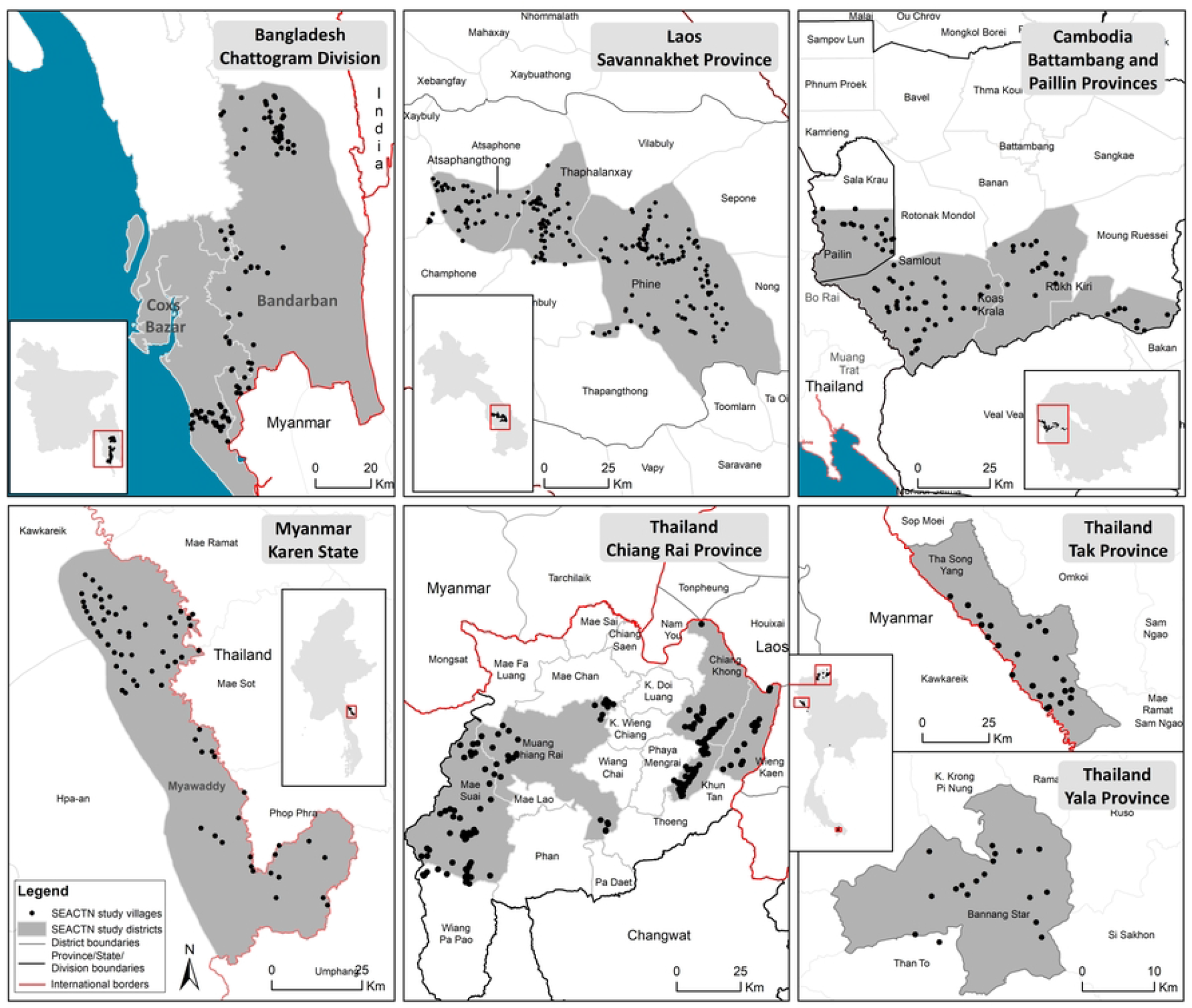
Locations of profiled villages within each study site of the South and Southeast Asian Community-based Trials Network.

### Demographic details and health and socio-economic status indicators

Site-specific demographic and health and socio-economic status indicator data are shown in Table 1.

**Table 1.** Demographic and key health and socio-economic indicator data for rural villages in the South and Southeast Asian Community-based Trials Network by site. Estimates are presented as medians (first quartile, third quartile) and n (%) unless otherwise indicated. *Completion of at least five years’ schooling was used as an indicator of literacy given varying definitions of literacy between countries. ^§^A delivery attended by a skilled birth attendant. ^†^A child older than one year who has received the following vaccines: Bacille Calmette–Guérin; three doses of diphtheria, tetanus, and pertussis; three doses of measles, and polio. ^‡^Proportion of households across all villages at each site which have either flush toilets or latrines. NA, not applicable; ND, data not collected or not available due to local restrictions.

| Country | Thailand |  |  | Laos | Cambodia | Myanmar | Bangladesh |
| --- | --- | --- | --- | --- | --- | --- | --- |
| Site (number of villages profiled) | Chiang Rai province (186) | Tak province (37) | Yala province (24) | Savannakhet province (194) | Battambang and Pailin provinces (81) | Karen state (35) | Chattogram division (130) |
| Number of key informants per village, n | 1 | 1 | 1 | 1 | 1 | 2 | 2–3 |
| **Demographic details** |  |  |  |  |  |  |  |
| Population per village, n | 555 (422, 769) | 902 (501, 1245) | 345 (238, 621) | 750 (486, 1135) | 639 (417, 1127) | 676 (340, 1021) | 526 (240, 1366) |
| Households per village, n | 180 (129, 234) | 240 (122, 372) | 101 (78, 147) | ND | 169 (100, 275) | 129 (64, 215) | 121 (57, 267) |
| Main languages at site, n | 10 | 4 | 2 | 4 | 1 | 8 | 8 |
| Main religions at site, n | 3 | 4 | 2 | 3 | 2 | 6 | 3 |
| **Health and socio-economic indicators** |  |  |  |  |  |  |  |
| Number of villages with a health facility within 30 min walk (%) | 105 (56.5) | 35 (94.6) | 15 (62.5) | 55 (28.4) | 61 (75.3) | 35 (100) | 114 (87.7) |
| Longest travel time to closest health facility by motor vehicle if none within 30 min walk (min) | 60 | 30 | 15 | ND | 90 | NA | 80 |
| % adults with ≥5 years schooling* | 75 (65, 100) | 60 (50, 85) | 60 (50, 70) | ND | 100 (90, 100) | 50 (70, 75) | 70 (60, 80) |
| Male | 75 (60, 90) | 50 (50, 90) | 50 (45, 55) |  | 100 (75, 100) | 60 (40, 70) | 65 (50, 75) |
| Female | 70 (55, 80) | 60 (50, 90) | 70 (65, 75) |  | 100 (55, 100) | 70 (55, 80) | 65 (45, 80) |
| % attended deliveries<sup>§</sup> | 100 (100, 100) | 10 (5, 27.5) | ND | ND | 100 (100, 100) | 80 (47.5, 95) | 60 (25, 85) |
| % fully-immunised children<sup>†</sup> | 100 (100, 100) | 100 (95, 100) | 67.5 (58.8, 75) | ND | 100 (100, 100) | 75 (85, 95) | 100 (100, 100) |
| % latrine coverage<sup>‡</sup> | 97.3 | 83.1 | 92.3 | ND | 86.7 | 84.8 | 67.5 |

In the profiled SEACTN villages, the median village population for any given site ranged between 345 and 902 people. There was a high level of cultural, religious, and linguistic diversity with several languages being spoken and religions practised at all sites, except in Cambodia and Yala province of Thailand which were more homogenous.

In general, the median percentage of adults with five years’ schooling was no greater than 75% except in Cambodia where this figure was 100%. There was little difference in educational attainment between genders except in Yala province, Thailand, where a median of 70% of females had achieved this level of education compared to 50% of males.

Except for Chiang Rai and Yala provinces in Thailand, latrine coverage did not exceed the 90% threshold generally accepted as the coverage required to have a positive impact on the health of the community.[16] Latrine coverage was particularly low in Bangladesh at 67.5%.

Vaccination coverage was low in Myanmar and Yala (median coverage of 75% and 67.5%, respectively) relative to other sites which had near-perfect coverage.

### Access to health services

The percentage of villages with access to a health facility within 30 minutes’ walk was highly variable, ranging from 28.4% in Laos to 100% in Myanmar. The travel times to the nearest health facility for villages with no facility within 30 minutes’ walk ranged between 15–90 minutes by motor vehicle. The median percentage of deliveries attended by skilled birth attendants was also markedly lower in Bangladesh (60%) compared to other sites.

As might be expected, few villages at all sites had access to doctors; the percentage of villages which had a doctor on site or within 30 minutes’ walk ranged from 4.2% in Yala to 20.0% in Myanmar (Figure 3a). By far the commonest types of health worker in the studied villages were primary health centre (PHC) workers/nurses and village health workers/village malaria workers (VHWs/VMWs). Interestingly, traditional healers were not found in most villages across all sites with the exception of Yala, where 70.8% of villages have them. All villages had access to a healthcare worker. Nevertheless, it should be borne in mind that many healthcare providers are informal, with little or no official training. These informal healthcare providers were found in 63.1% and 77.1% of villages in Bangladesh and Myanmar, respectively and do not include traditional healers or traditional birth attendants; however, in all of the villages where they were present, there was also at least one type of formal healthcare provider.

**Figure 3.**
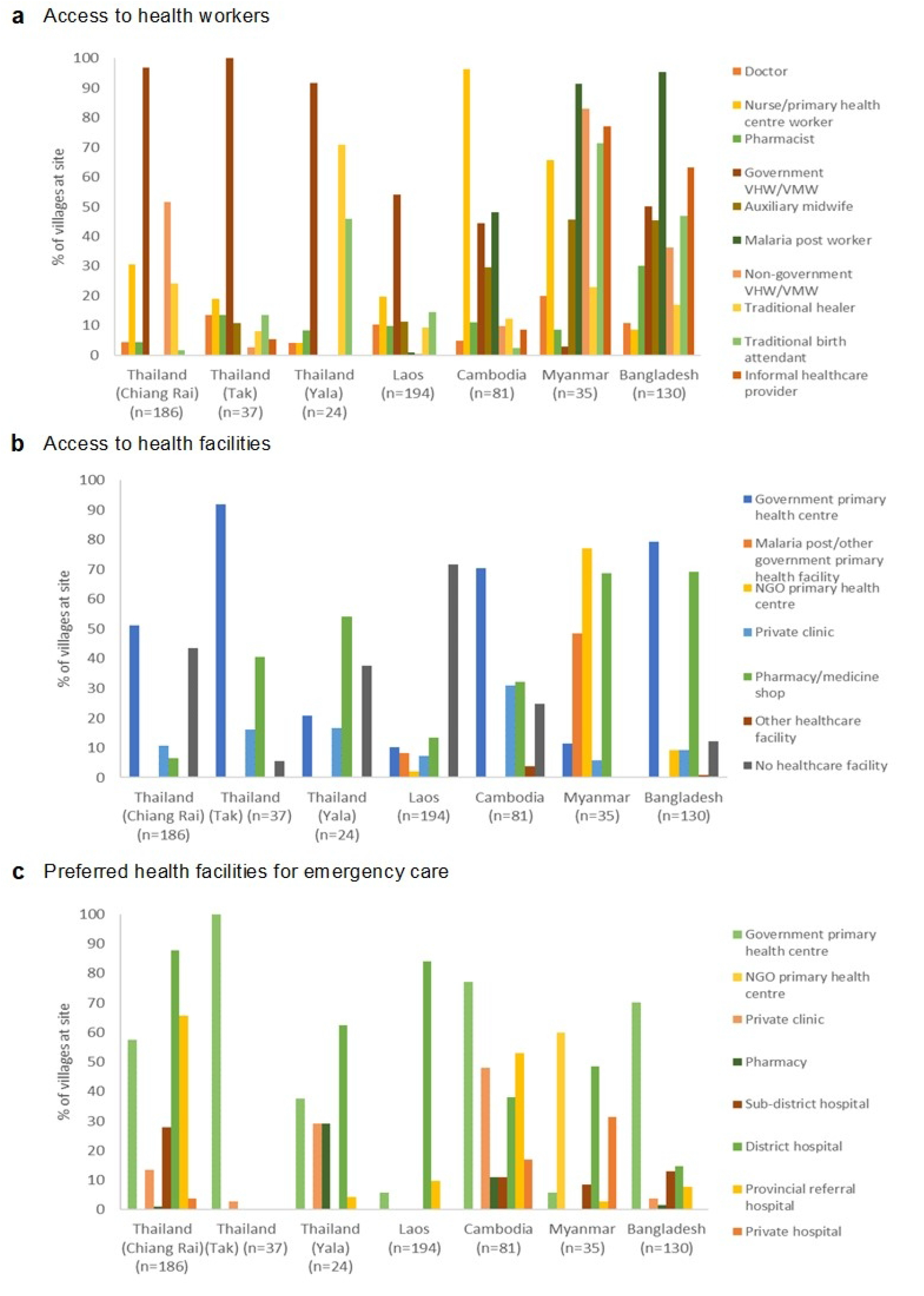
Percentages of villages in South and Southeast Asian Community-based Trials Network Rural Febrile Illness project study sites with access to different types of (a) healthcare worker and (b) healthcare facility. Percentages of villages and the types of healthcare facilities at which patients perceived as being severely ill from those villages would seek care are shown in (c). Access was defined as having the healthcare worker or facility in the village or within 30 minutes’ walk. NGO, non-governmental organisation; VHW/VMW, village health worker/village malaria worker.

It should be of little surprise given the above findings that most facility-based healthcare in the profiled villages was delivered at PHCs (Figure 3b), and that where they are not, such as in Yala and Laos where there are few PHCs, the burden of primary healthcare delivery falls on VHWs/VMWs. This is most important in Laos, where 71.6% of villages had no healthcare facility, including pharmacies or medicine shops, within 30 minutes’ walk. Notably also, in Myanmar 77.1% of villages were served by clinics set up by non-governmental organisations (NGOs) vs. 11.4% served by government PHCs. It is also important to note the relatively easy access to pharmacies or medicine shops at all sites except Chiang Rai and Laos, especially since medicines are generally available over the counter.

As shown also in Figure 3b, there are no secondary care facilities able to care for severely ill patients within 30 minutes’ walk of any profiled village, with all easily accessible healthcare facilities being designed for the provision of basic primary care and/or diagnosis and treatment of uncomplicated malaria, in the case of malaria posts. It is, therefore, not unexpected that patients who perceive themselves as being severely unwell would generally primarily seek care from first-level referral hospitals i.e., those at the sub-district and district levels, with the exception of Yala, Cambodia, and Bangladesh (Figure 3c). At these three sites, the commonest facility to which severely unwell patients would present remains the PHC; this was a preferred option for emergency care in 37.5%, 77%, and 70% of villages, respectively. However, the large role of private sector facilities, such as private hospitals and clinics, in treating severely ill patients in Cambodia, Myanmar, and Yala should also be noted; at these sites, such facilities are a preferred option for emergency care in 65.0%, 31.4%, and 29.2% of villages, respectively.

### Access to public utilities and education

#### Water and sanitation

Thai villages had the best access to improved drinking water sources, with the majority of villages having access to piped water available for at least most of the day (Table 2). Additionally, over half of the villages studied in the Thai provinces of Chiang Rai and Yala were able to obtain bottled water and/or filtered or distilled water via dispensers; in contrast, fewer than 10% of Cambodian and Lao villages were supplied with piped water, with the principal water sources being wells and boreholes (Figure 4a). Overall, however, a higher percentage of villages had access to improved than unimproved drinking water sources, and the majority of villages without piped water had their water sources located within the village (Figure 4a).

**Table 2.**
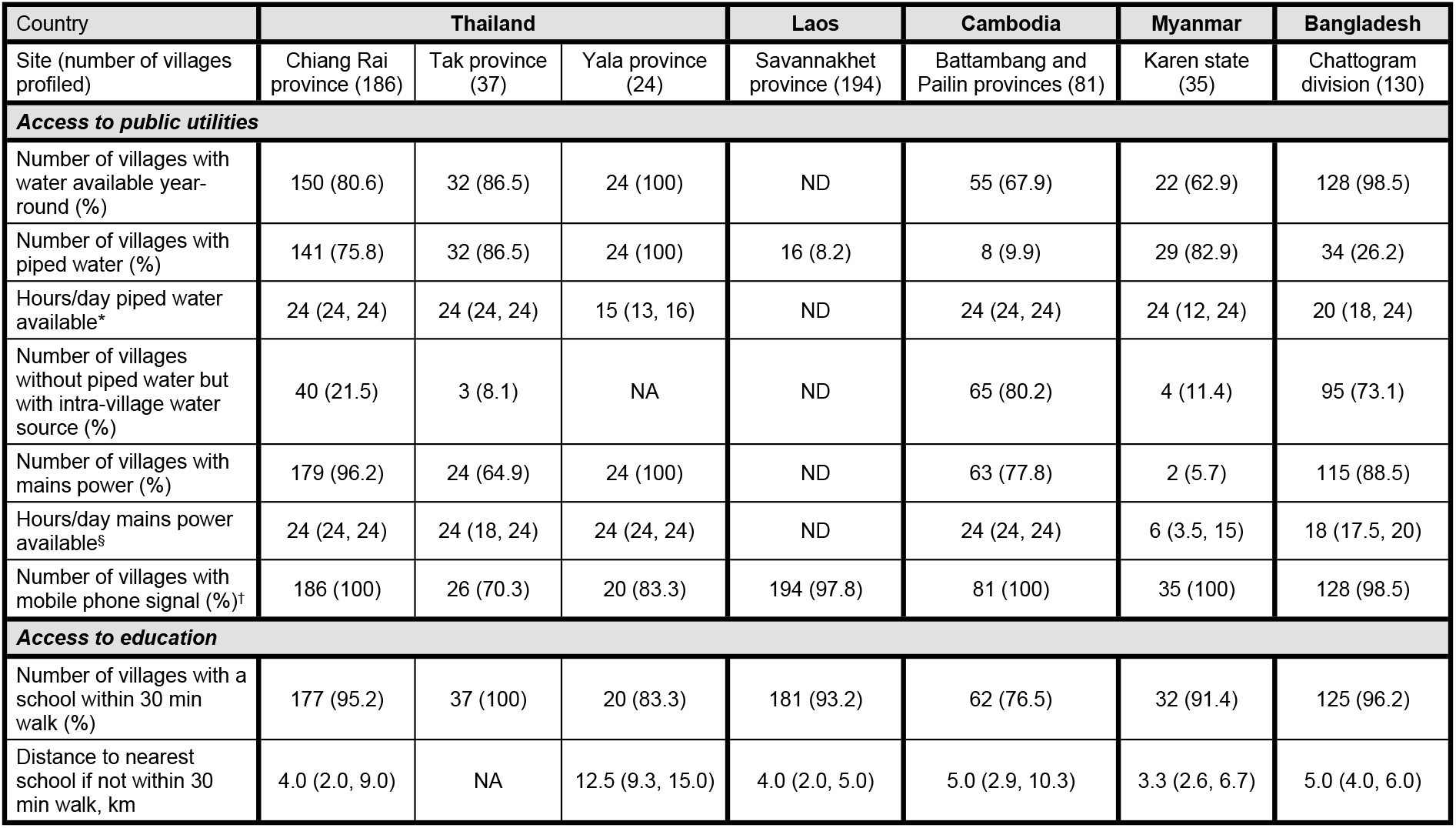
Access to public utilities and education for rural villages in the South and Southeast Asian Community-based Trials Network by site. The number of hours per day piped water and mains power were available, and distance to nearest school if not within 30 minutes’ walk, are presented as median (first quartile, third quartile). *For villages with piped water. ^§^For villages with mains power. ^†^Defined as mobile phone signal present at least some of the time. NA, not applicable; ND, data not collected or available due to local restrictions.

**Figure 4.**
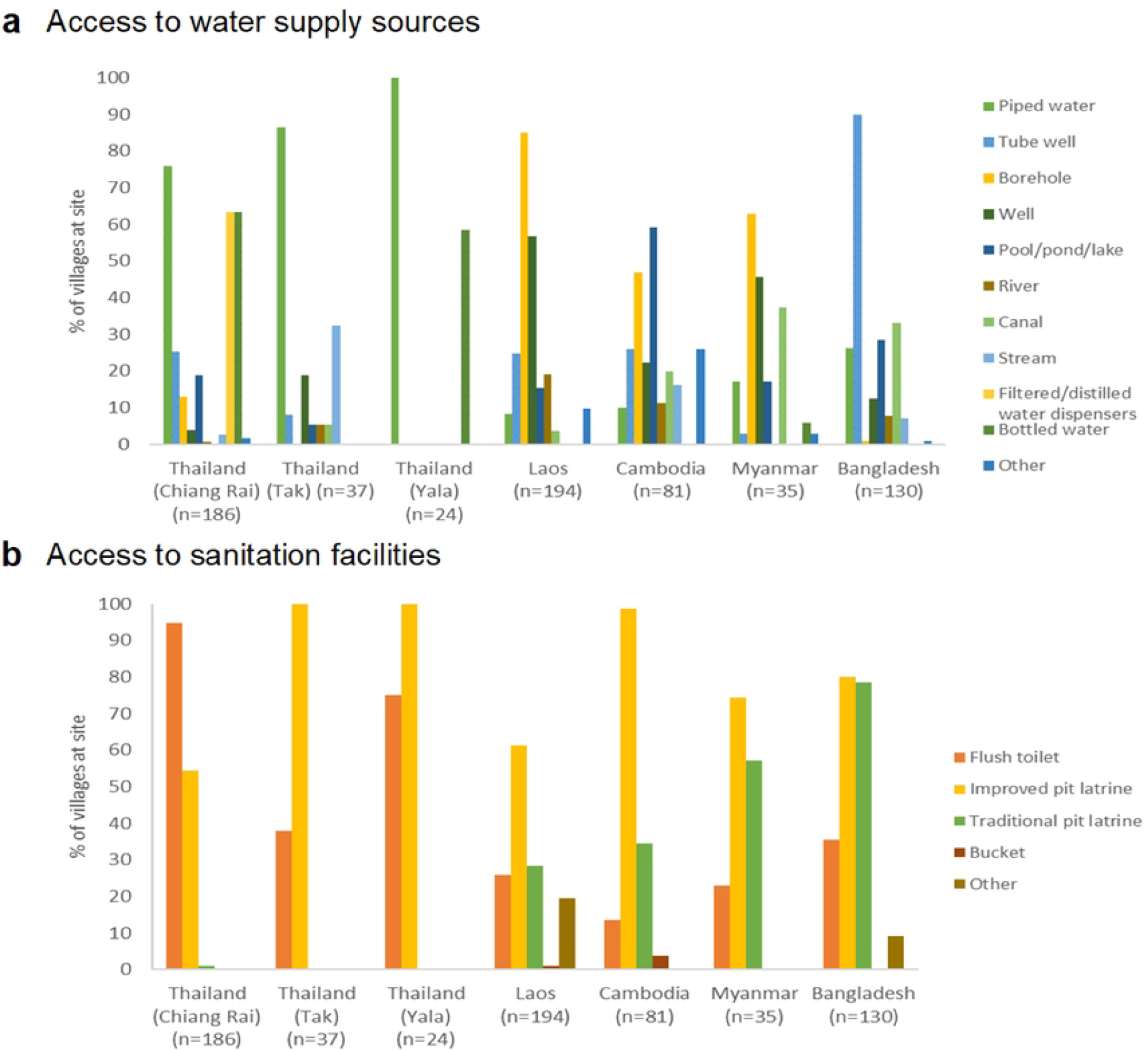
Percentages of villages in South and Southeast Asian Community-based Trials Network Rural Febrile Illness project study sites with access to different types of (a) water supply sources and (b) sanitation facilities. Access was defined as having the water source or sanitation facility in the village or within 30 minutes’ walk.

Unsurprisingly, the quality of sanitation facilities at each site mirrored the availability of piped water, with the percentage of villages with access to flush toilets being the highest in Chiang Rai (94.6%), and the lowest in Cambodia (13.6%) (Figure 4b). Overall, most villages relied on improved pit latrines (Figure 4b). However, data on whether these were shared/public vs. private were not collected, although it is likely that at least some were shared facilities, especially in Bangladesh where 6.1% of villages were noted to have non-functioning latrines.

#### Electricity and light sources, and mobile phone coverage

With the exception of Myanmar, over 60% of villages at each site had access to mains electricity, with the highest penetration being in Yala (100%). Only 5.2% of villages at the Myanmar site were supplied with mains power; at this site, electricity was mainly provided by via generators or solar panels (Figure 5a). However, even in villages with mains power, supply was not always reliable, with the median number of hours per day ranging between six in Myanmar to 24 in Thailand (Table 2).

**Figure 5.**
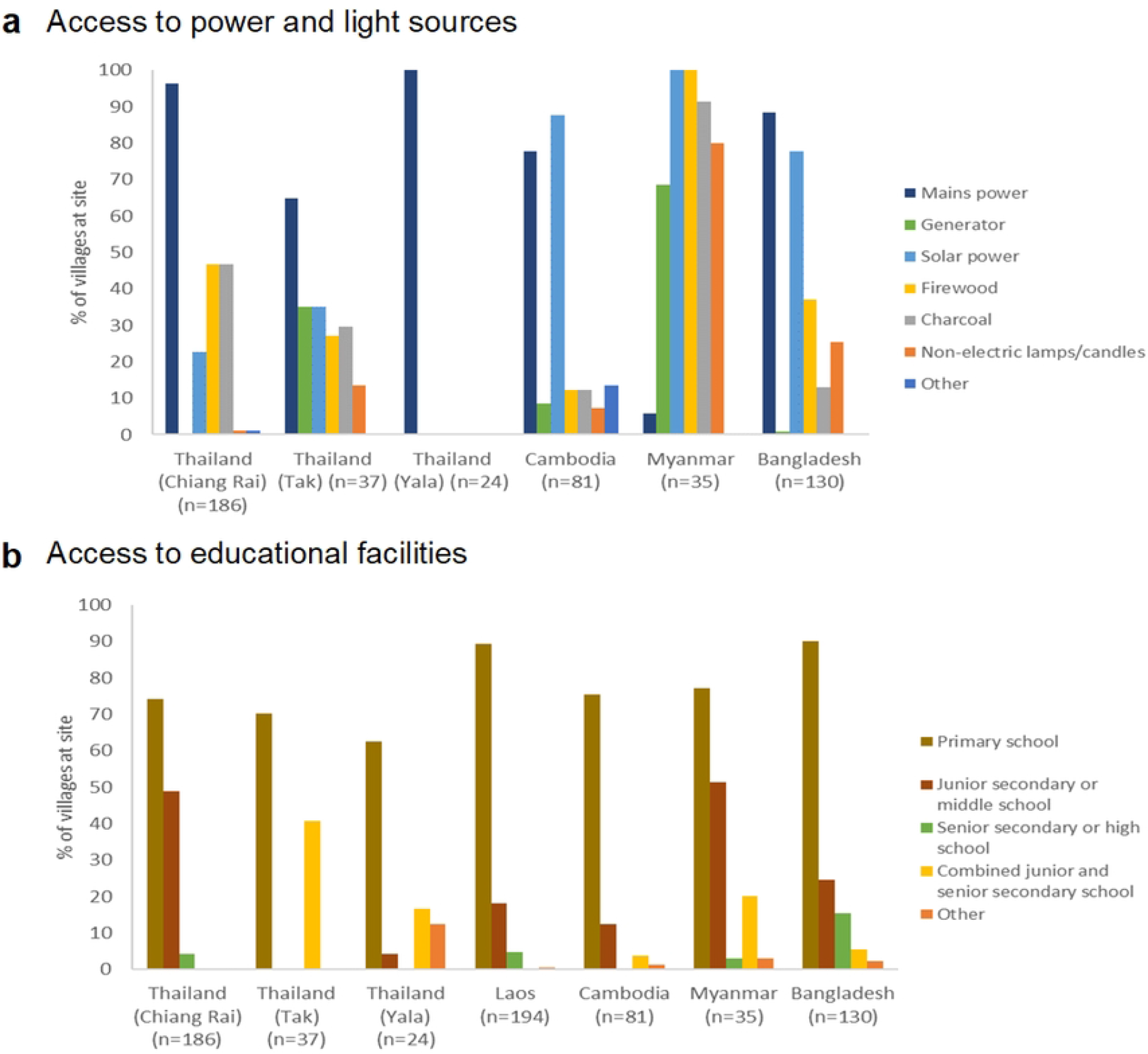
Percentages of villages in South and Southeast Asian Community-based Trials Network Rural Febrile Illness project study sites with access to different types of (a) power and light sources and (b) educational facilities. Access to power and light sources was defined as having the power and light source in the village, while access to educational facilities was defined as having the facility in the village or within 30 minutes’ walk.

Regardless of source, electricity was generally available at the profiled villages even if not all households had reliable and affordable access to it, as evident by the continued use of firewood, charcoal, and candles (Figure 5a). The availability of electricity supports the use of mobile phones, evidenced by the widespread signal coverage across all sites (Table 2).

#### Educational facilities

Across all sites, the majority of villages had a primary school within 30 minutes’ walk (Figure 5b), with an even higher percentage having any type of educational facility within this distance (Table 2). The site with the lowest access to educational facilities was Yala, where 62.5% of villages had a primary school within 30 minutes’ walk (vs. 90% in Bangladesh). Furthermore, for villages in Yala with no school within this travel time, the median distance to the nearest school was longest compared to similar villages at all sites at 12.5 km (vs. ≤5 km at the other sites) (Table 2).

## Discussion

This study profiled 687 villages located in five South and Southeast Asian low-income and middle-income countries (LMICs) at different stages of development, revealing very different characteristics, especially in terms of access to services and public utilities. In line with this, therefore, there was considerable variation between sites in terms of health and socio-economic indicators.

Nevertheless, several common themes were evident, the first being the obvious ethno-cultural and linguistic diversity that exists even within villages at the same site. The second is their reliance on PHCs (and analogous NGO primary health clinics), and VHWs/VMWs, as the backbone of primary healthcare provision. The latter are particularly common in Thailand, Laos, Bangladesh, and Cambodia where they have been a longstanding feature of village healthcare over the past decades. However, severely ill patients would generally travel out of their villages to seek care at first-level referral hospitals and private sector facilities in towns, bypassing village-level health services. Thirdly, while most villages were proximate to a primary school, the level of education was generally low. Fourthly, mobile phone coverage and availability of electricity was generally high.

Several findings require further explanation. Firstly, the low vaccine coverage in Myanmar and Yala may partially be explained by these locations being conflict zones,[10, 17] with an additional layer of mistrust of the government and, by extension, government programmes by the majority Malay ethnic group in Yala.[17] Furthermore, the low coverage of villages by public sector PHCs in Myanmar is due to the traditionally meagre government expenditure on healthcare,[18] encouraging a parallel system of primary health clinics set up by NGOs to meet demand. This is especially so in Karen State where control by the central government has been patchy.[19] Lastly, while not all villages were supplied by electricity through power grids and, even in those that were, the number of hours per day that such supply is available is limited yet mobile phone coverage was high across all sites. This is due to innovative solutions having been devised to overcome the lack of private household electricity supply, such as grocery shops installing shared solar-powered mobile phone charging points in villages in Myanmar.[20]

The principal strength of this study is its focus on key health and socio-economic indicators, data on which public health and other policymakers, as well as researchers, can use for planning purposes. Furthermore, a pragmatic, low-cost, semi-desk-based methodology using key informants was chosen. This approach allowed reasonable estimates of the key indicators in the absence of easily accessible official information for all domains, in addition to the difficulties posed by the remote locations of many villages and travel restrictions for security reasons to some e.g., those in the Chattogram Hill Tracts in Bangladesh. It also has the benefit of allowing high-quality spatially granular data to be collected, permitting detailed comparisons to be made. The key informants were selected by staff of SEACTN country partner organisations to ensure a high degree of first-hand knowledge of the prevailing conditions in their respective villages, as is best practice.[21] This approach is well-suited to the objectives of this study, as it is appropriate if general, descriptive information is sufficient to guide decision-making, generate suggestions and recommendations, and assist in the design of more comprehensive quantitative research.[22] Key informant interviews are also appropriate for when quantitative data collected through other methods need to be interpreted,[22] which is exactly the situation with SEACTN where the data from the observational epidemiological studies being conducted at these sites need to be viewed through a contextual prism.[23]

Nevertheless, while the results provide an idea of the characteristics of villages at the individual sites, they may not be generalisable to all rural villages within their respective countries. This is amply demonstrated by heterogeneity of the three Thai sites, despite all being located in remote areas bordering neighbouring countries; there are likely to be many more differences between these borderland villages and those in the more developed central plains. In the case of Cambodia, the relative ethno-linguistic homogeneity and high elementary educational attainment of the villages profiled for this study would not likely be found in villages in the northeast of the country bordering Laos, where more languages are spoken and literacy rates lower.[24] The selected villages at the Myanmar site had better road and telecommunication links than others due to the logistical and security considerations previously described. This is likely to have resulted in better access to water and sanitation, health services, and educational facilities, which may not be representative of the typical conditions elsewhere in Karen state or Myanmar in general. For example, there is an international hospital in Shwe Kokko close to several villages profiled at the Myanmar site, whereas no such facilities exist deeper in the interior of Karen state. Also, the higher immunisation coverage in these villages compared to the very low or negligible figures reported for villages elsewhere in Karen state,[25] is likely due to the ability of residents to access the more advanced health services in Thailand, including preventative care, as mentioned earlier.

Additionally, there are several well-known limitations of the key informant interview approach that should be borne in mind when interpreting the results of this study. Chief of these is the potential for bias since it was not possible to confirm the appropriateness of informant selection; for example, more qualified but less visible or prominent informants may not have been selected.[26] It is also possible that, due to their prominence, the socio-demographic strata from which key informants are drawn may not be similar to that of the majority of village dwellers, in particular those from the most marginalised groups, leading to a perhaps less than comprehensive understanding of their villages. This should be taken into account when interpreting the results, since their potentially differing perspectives may have caused them to overestimate their responses to questions on health and socio-economic indicators. It is particularly important to consider this effect at the Myanmar site since, added to that of the relatively better access to services of the profiled villages at this site, it may have resulted in results which are less representative of more typical villages there. Another source of bias that should be considered is interviewer bias, although this was mitigated by using a data collection tool with questions to be read out verbatim by the interviewer, as well as having many questions with response options or which required numerical responses. However, the questions which required quantitative responses were in themselves a weakness, since key informant interviews are best suited to qualitative studies because they provide only a very restricted basis for quantification.[22] It is also difficult to verify the validity of the findings because only a very small number of informants were selected per village, certainly fewer than the ideal figure of 15–35.[26] A final limitation of this study is that it while it may indicate the presence or absence of particular facilities or services, it gives no indication of their quality.[27]

There are two obvious priorities for further research. The first is to flesh out the findings with the results of other studies examining broadly similar themes, such as the SEACTN household health survey.[28] In addition, more detailed site-specific analyses triangulating the findings of this study with geospatial epidemiological work analysing accessibility to health and other services will be extremely informative. Such efforts will allow further layers of contextual understanding to be built on the foundations established by this study. Secondly, this study has uncovered the major public health and socio-economic issues important to each site, for example the low vaccination rates in Yala province, the low percentage of births attended by skilled attendants in Bangladesh, and the globally poor levels of educational attainment despite relatively easy physical access to primary schools. Future studies should explore the reasons behind such problems in the light of the other contextual findings, with a view to developing potential interventions to solve these issues.

## Conclusion

This study provides a rudimentary descriptive overview of the characteristics of under-served and under-studied villages in rural South and Southeast Asia, focusing on key health and socio-economic indicators and the availability and accessibility of amenities and public services essential to good health, such as water and sanitation. Notwithstanding its weaknesses, the results illustrate the wide diversity in the region and this should be considered in public health research and policymaking.

## Funding

This research was funded in whole, by the Wellcome Trust [220211/Z/20/Z and 215604/Z/19/Z]. RC was also funded by the UK Government through a Commonwealth Scholarship, the Royal Australasian College of Physicians through the Bushell Travelling Fellowship in Medicine or the Allied Sciences, and the Rotary Foundation through a Global Grant Scholarship.

The funders had no role in study design, data collection, data analysis, data interpretation, or writing of the manuscript. All authors had full access to the data and had final responsibility for the decision to submit the manuscript for publication. For the purpose of open access, the authors have applied a CC BY public copyright license to any Author Accepted Manuscript version arising from this submission.

## Competing interests

The authors declare no competing interests.

## Data availability

All data relevant to the study are included in the article or available upon reasonable request from the corresponding author.

## Notes

### Competing Interest Statement

The authors have declared no competing interest.

### Author Declarations

Ethical approval was not required for this study.

